# Caffeine and Sleep in East Tennessee students

**DOI:** 10.1101/2022.11.04.22281900

**Authors:** Viraj Brahmbhatt

## Abstract

Caffeine is a psychoactive stimulant that works on the central nervous system and is widely consumed for its ability to increase alertness. The well-known effects of caffeine consumption include increasing alertness and focus. In addition, this caffeine consumption is well known for its ability to interrupt sleep. In previous studies investigating the prevalence of caffeine consumption, students indicated that caffeine consumption was a prevalent behavior sought after to improve academic and athletic performance in the college students studied (Mahoney et al., 2019). Current American Academy of Pediatrics guidelines for caffeine consumption of 100 mg/day for those under 18 seem optimistic given the prevalence of caffeine consumption indicated in current literature. Current studies in medical students in the Middle East have indicated high levels of caffeine consumption and lacking sleep as well as elevated anxiety and depression symptoms. This study aims to bridge the gap on caffeine consumption, sleep, and associated behaviors in American students in the East Tennessee area. The method utilized a survey distributed through the REDCap platform. The survey was constructed using the Pittsburgh Sleep Quality Index (PSQI) as well as an original caffeine consumption portion of the survey. An open response section was provided so that respondents may be able to provide specific factors that may have contributed to their loss of sleep or increased caffeine intake. The final sample included 97 responses from high school and college students between the ages of 14 and 45, of which 57 were deemed fully completed and able to be analyzed. Analysis revealed that there was a correlation between decreased sleep and increased caffeine intake. 63.2% of respondents indicated caffeine consumption. For the individuals 18 and under, 75% of respondents indicated caffeine consumption over the healthy guidelines indicated by the American Academy of Pediatrics. Additionally, in the free response section, 27% of respondents indicated that they were unable to sleep due to stress brought on by school or homework, while 15% of respondents indicated that they had been able to get sufficient sleep because of stress. This study offers insight into the higher than recognized caffeine usage within students. Additionally, sleep levels were generally below the recommended guidelines. This data prompts further studies looking into adolescent mental health, associated with the lacking sleep and high caffeine levels, as well as prompts schools to potentially offer additional resources to combat the mental health detriment that may be suffered as a result of lacking sleep and excessive caffeine intake.

## Introduction

Caffeine is a psychoactive stimulant that works on the central nervous system and is widely consumed for its ability to increase alertness. Embedded in cultures around the world, caffeine is found in substances ranging from coffee and tea to chocolate and sodas. Caffeine intake produces a physical reaction of blood vessel constriction and blood pressure elevation, which is linked to the feelings of alertness associated with the substance. Though regarded as a non-addictive substance, caffeine use may lead to “abstinence syndrome”, or the phenomena of experiencing negative feelings which may range from headaches and irritability to a depressed mood and anxiety (Angelats et al., 2017). In addition to this, caffeine consumption, in the form of energy drinks/shots, was linked with lesser perceived risk in drug use in adolescents, according to a 2018 Drug Alcohol Dependence publication (Jackson & Leal, 2018). The results of this study were meant to alert for the correlation of increased drug use with high caffeine consumption in adolescents.

Beyond the symptoms and possible drug-use correlations, caffeine can be detrimental to sleep-wake cycles. A publication from the Current Opinion in Physiology journal discussed the importance of sleep as it could be seen in that it is the period in which brain maturation as well as emotional and cognitive development occurs. During periods of development – notably adolescence (13 - mid 20’s)– the importance of sleep is monumental (Fontanellaz-Castiglione, C. et al, 2020). The lack of sleep can lead to impaired cognitive functionality as well as attention lapses, and decreased vigilance, attention, and speed of psychomotor responses (Goel et al, 2009). In fact, a study conducted on Saudi Arabian Medical Students reported the finding that less sleep was linked with much greater rates of stress, anxiety, and depression symptoms (Al-Khani et al, 2019). The detriment of sleep loss may even be seen on a physiological level. A 2018 Urilla et al publication commented on the visible correlation between sleep loss and lesser gray matter volume. This publication presents the development of physiological markers resulting from detrimental, disrupted sleeping patterns (Urrila et al., 2017).

Sleep deprivation is seen as a part of the initiation of caffeine intake. This being said, caffeine intake may lead to sleep interruptions. A 2018 publication from the Risk Management and Healthcare Policy commented that this cycle of sleep deprivation can be seen as contributing to increased caffeine consumption. This cycle can be summarized in caffeine interrupting sleep and subsequent use of caffeine to compensate for the lack of sleep (O’Callaghan et al, 2018). The observed effect is a compounding caffeine increase as well as a compounded effect of sleep deprivation and caffeine intake.

The article “Adolescents Drink Too Much Caffeine” from Medical News Today cited that 96% of teenagers occasionally consume caffeinated beverages, with 83.2% of teenagers regularly consuming caffeinated beverages (Brazier, 2016). USDA guidelines quoted average caffeine intake for adolescents at under 100 mg/day, affirming that this level of intake is well under the healthy threshold of 400 mg/day (US Department of Health and Human Services and US department of Agriculture, 2015). (Seidenberg, 2018l). For individuals over the age of 18 (ages 19-30), the estimated caffeine consumption was noticeably greater. The Dietary Guidelines for Americans quote current levels of caffeine consumption at between 110 and 260 mg daily (US Department of Health and Human Services and US Department of Agriculture, 2015).

It is also important to consider the motivations behind the consumption alongside the rate of consumption. Notably the positive effects of alertness, elevated focus, and mood elevation associated with the substance (Choi, 2020). Students may chase these qualities to improve academic performance or to compensate for decreased performance. From the perspective of the student, increased alertness and focus may provide the means to obtain the desired academic performance. A similar effect may be extrapolated to athletes and athletic consumption of caffeine. A 2019 publication from the *Nutrients* Journal commented on the noticeable increase in power produced, weight lifted, and sprint performance for athletes consuming supplemental caffeine in comparison to athletes consuming the placebo (Mielgo-Ayuso et al., 2019).

The intake guidelines of no more than 400mg/day may be seen as optimistic when analyzing the caffeine composition of many common beverages. Coffee drinks may contain upwards 235 mg (small coffee) while energy drinks and sodas may contain caffeine ranging from 30 to 200mg. Though it may seem as even these substances contain caffeine levels well below the 400mg threshold, caffeine intake for adolescents is dictated by separate guidelines due to the differences in caffeine processing in adolescents (Seidenberg, 2018). The American Academy of Pediatrics quoted these reassessed guidelines as 100mg as a safe upper limit for caffeine intake in adolescents. These guidelines were presented alongside the guidelines for other groups in a 2017 Food Insight publication (Insight, 2017).

Though the current quoted rates of caffeine intake are low, this reported rate is likely lower than the true rate of consumption. In a 2011 study published in the Journal of Caffeine Research, the substance dependence criteria was analyzed in relation to caffeine intake for high school and college students. The study proposed that 35% of students surveyed met criteria for caffeine dependence. This rate is quite high given the optimism shown in USDA guidelines for caffeine use, suggesting there is greater adolescent caffeine consumption than previously suggested (Striley et al., 2011).

## Literature Review

To understand the high rates of adolescent caffeine intake, it is important to analyze the contributors to caffeine intake. Caffeine has become a compound associated with cultural norms, acclaimed for its ability to increase focus (as well as alertness and athletic performance), which is a quality sought by students (Is Caffeine Good or Bad for Studying?, 2020). However, this caffeine intake can become the cause of a sleep deficit. A publication from the American Journal of Public Health criticized current early school start times for being an unhealthy practice in relation to sleep duration and consequent mental health detriment in adolescents (Paksarian et al., 2015 ; Peltz et al., 2017). A 2016 publication from the Journal of caffeine research reported that caffeine intake in students was correlated with disturbances in healthy sleep schedules - these disturbances were more pronounced in energy drink consumers. Additionally, regular energy drink consumption was correlated with poor sleeping patterns, mental illness, and substance use risk behaviors (Kelly & Prichard, 2016).

Current studies discuss surrounding factors of adolescent lifestyles - such as electronic use, exercise, etc. – in contributing to inadequate sleep in adolescents. There are even studies that individually observe the connection between early school start times and inadequate sleep as well as studies that found connections between inadequate sleep and subsequent caffeine intake (Jahrami et al., 2020). Notably, a 2019 publication on the correlations of sleep and mental health in Saudi Arabian medical students (Al-Khani et al). The referenced study brings up relevant concerns relating to the unhealthy sleeping habits and caffeine consumption habits that seemed to pervade the institution. This study prompts the question of the prevalence of similar habits in students of different levels as well as students in other countries-specifically the United States. There is a noticeable gap in the existing literature in terms of caffeine consumption and correlation to sleep in terms of students in East Tennessee.

A 2009 publication from the Pediatrics journal published findings suggesting that there is caffeine use in adolescents resulting from inadequate sleep the night before. This particular publication looked into the role of electronic use and inadequate sleep (Calamaro et al, 2009). A major takeaway was that adolescents, generally, have inadequate sleep which can affect their academic performance. This notion was reflected in a 2009 The Journal of Biological and Medical Rhythm Research publication. This particular study discovered that sleeping patterns were worse in male adolescents (Tonetti et al, 2008). This evolving knowledge that adolescent sleeping patterns were inadequate led to research into other factors which may be impacting the sleeping schedules. Early school start times – school start times before 8 am – were seen as being contributory to inadequate sleep as well as poorer quality of sleep, as discussed in an article from the American Journal of Public health (Paksarian et al., 2015).

Though it may seem that periods of inadequate sleep and caffeine consumption may be a benign phenomena, there are significant long-term effects associated with the two. Sleep loss has been associated with the onset of obesity, diabetes, cardiovascular disease, anxiety, depression, and alcohol use (Colten & Altevogt, 2006). Sleep deprivation in students is particularly detrimental. A Stanford Medicine publication commented on the current state of sleep deprivation as an epidemic in students-specifically teens (Richter, 2015). A 2014 publication from the Nature and Science of Sleep journal discussed the specific dangers associated with sleep deprivation in students. Along with the decreased grades and learning efficiency, sleep deprivation is correlated with increased risk of motor vehicular accidents (Hershner & Chervin, 2014). This risk becomes especially relevant when one considers that automobile accidents are the greatest risk factors for adolescents and young adults (“Adolescent and young adult health”, 2021).

Additionally, high caffeine intake has been linked with the onset of anxiety, ulcers, irritability, headaches,dizziness, and weakness (“Caffeine”, 2020). Seeing as sleep deprivation and caffeine use are linked, the combined effects of the two are compounded and more extreme combined.

## Methods

This study was conducted using a self-assessment questionnaire in order to assess the correlation between caffeine use and school start times. The questionnaire was composed of two section:a sleep quality index and a caffeine consumption section. The sleep quality portion was adapted from the standardized Pittsburgh Sleep Quality Index (PSQI). The caffeine use questions were made by the principal investigator after consulting existing literature. A copy of the questions is presented in the Appendix section.

The caffeine consumption component of the survey looks into the amount of consumption as well as the feelings associated with the consumption. Throughout the survey, questions were adjusted from open responses to a set of multiple choice responses to help narrow answers. For example, when asking about sleep duration, a participant may have been able to write “at least 6 hours”. Though this answer is reasonable, it does not provide the same information as “between 6 and 7 hours”. This is because, indicating sleeping more than 6 hours includes 7, 8,9, etc. hours and is not as specific as “between 6 and 7 hours”. Having a clear response criteria through the multiple choice answer choices increases the answer validity as the participant responses are narrowed. Multiple choice segments were often redundant or included an open response possibility. The redundant nature of “wake up time” or “earliest activity” as well as school start time questions were asked to increase validity of results through repeated confirmation. These questions also brought about increased clarity on the sleeping habits of study participants. The open answer segment, including the “other, please indicate”, areas are intended to incorporate a qualitative criteria into the study and to address the possibility of overlooking unique contributory factors. For example, a participant may indicate they consume caffeine as a substance to boost athletic performance, yet they have not felt any disturbances to their sleep. This personalized response contributes to the understanding of caffeine consumption and sleeping habits in the participants studied.

The questionnaire was first sent to administration at schools (Name of Schools Redacted), gaining permission to proceed with the survey distribution. The schools’ administrations were asked to distribute the electronic surveys to the students through email. Before the Survey could be accessed, participants were asked to provide their consent through a form. If the participant indicated that they were under the age of 18, a necessary parental consent form was displayed. Once parent consent was gained, the students were asked to provide consent. Given that both parties provided consent, the study was able to proceed.

The questionnaire was designed to take less than 15 minutes and students were asked to take the questionnaire in a quiet area when they have plenty of time (at home if possible). At one site, the survey distribution was approved through consulting the deans of academic departments and getting their permission to proceed. Collecting these permission letters from each site was an essential step of the Institutional Review Board (IRB) approval process that was deemed necessary as the study was categorized as human subjects research.

The data from the questionnaires was analyzed to determine any correlation present between school start times and caffeine intake. The questions about sleep will be used to contextualize the study findings in the importance of sleep and the health detriment of lacking sleep. A linear regression analysis was used to determine a general correlation between the variables of sleep and caffeine intake as well as caffeine intake and age. It is inferred that increased caffeine use should see a correlation to decreased sleep and that increase in age of participants should correlate with increased caffeine intake. The study aimed to contextualize the correlation amongst the variables of sleep, caffeine intake, and age.

## Results

The survey was distributed to a local highschool (High School A) and a local university (University A). Within the university, the survey was asked to be distributed by the deans of respective schools; however, only the dean of the school of nursing replied and this was where the survey data (for University A) was gathered from. For High School A, surveyors came from all highschool grade levels as well as ages. In total, the survey received 97 responses, of which 57 were completely finished. This entire was then sorted into categories prior to analysis. The primary groups were those 18 and under (the group created with the highschool students) and those above 18 (the college students). Aside from these groups, the entire group was also sorted and analyzed based on the age of the respondent. Within this cohort, 36.8% (21 responses) of responses indicated they did not consume caffeine, while 63.2% (36 responses) of responses indicated that they did in fact regularly consume caffeine. Of the cohort surveyed, only 18% indicated sleep that was considered sufficient by the Center for Disease Control (CDC) guidelines.

## Discussion

The study utilized a survey which attempted to collect information regarding caffeine consumption and quality of sleep. Upon analyzing the 57 fully completed responses for correlation among variables, certain trends were noticeable. This study took on a similar approach to the Jahrami et al. 2020 publication detailing the associations between caffeine intake and physical/mental health status (Jahrami et al., 2020). This being said, this study took on a different sample than the one detailed in the Jahrami et al. study. One specific correlation studied was the association between age and caffeine intake. Initially it was hypothesized that due to the dependence effect of caffeine, older ages would consume greater caffeine than younger ages. The trend for caffeine and age does not display significance. The trendline data for the correlation between age and caffeine consumption displays very little correlation. This refutes the initial hypothesis. This being said, there can be further analysis done through comparing caffeine consumption in the oldest and youngest age groups. This difference (shown in Figure 8.) speaks on the onset of caffeine consumption and the phenomena of an increased prevalence of caffeine consumption in older individuals as opposed to those who are younger.

**Figure 1.**
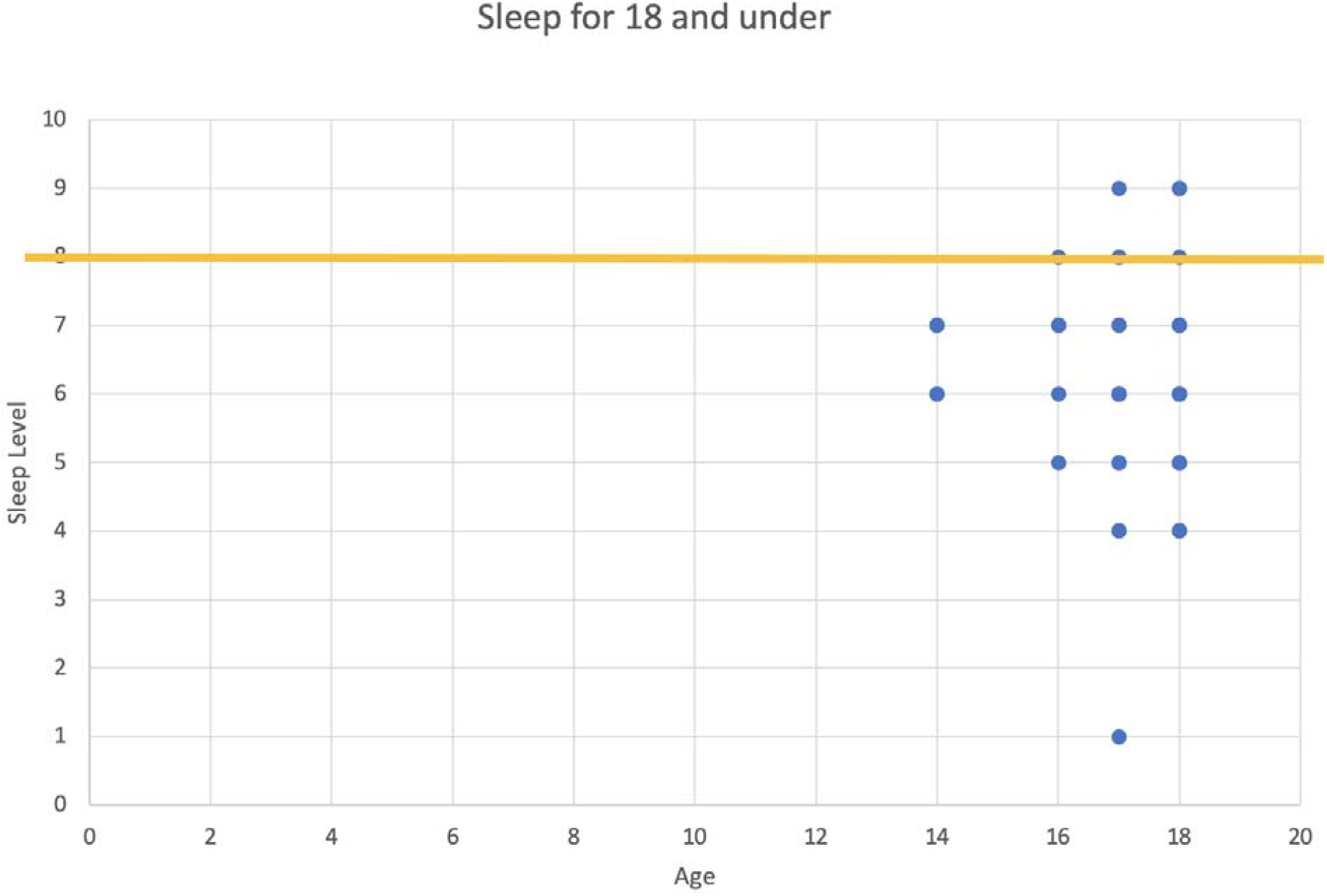
Figure 1. Displays the self-assessed sleep as compared to the CDC recommended average sleep (shown through the yellow line). The vast majority of surveyed individuals fell below the sleep recommendation. Given that this data is shown for both caffeine and non caffeine drinking individuals, it is to be inferred that the students are not getting enough sleep.

**Figure 2.**
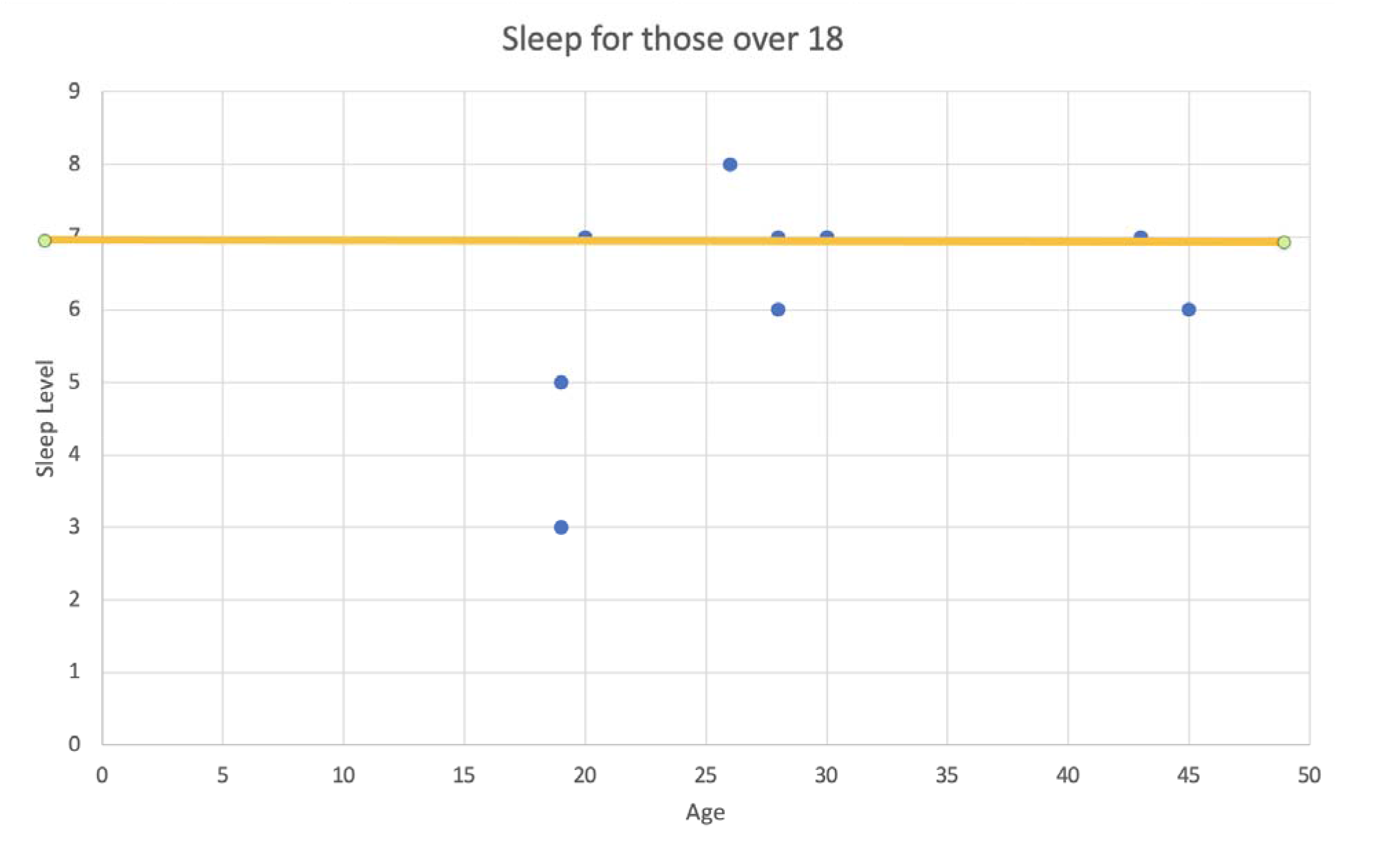
Figure 2. Displays an improvement over the sleep observed in the under 18 group. Still, there are a significant number of individuals that do not receive sufficient sleep as per CDC guidelines. For individuals over 18, sleep recommendations are more liberal as development during sleep has decreased. Given this information, it may be helpful to look into the sleep interruptions and how these change per evolving age groups.

**Figure 3.**
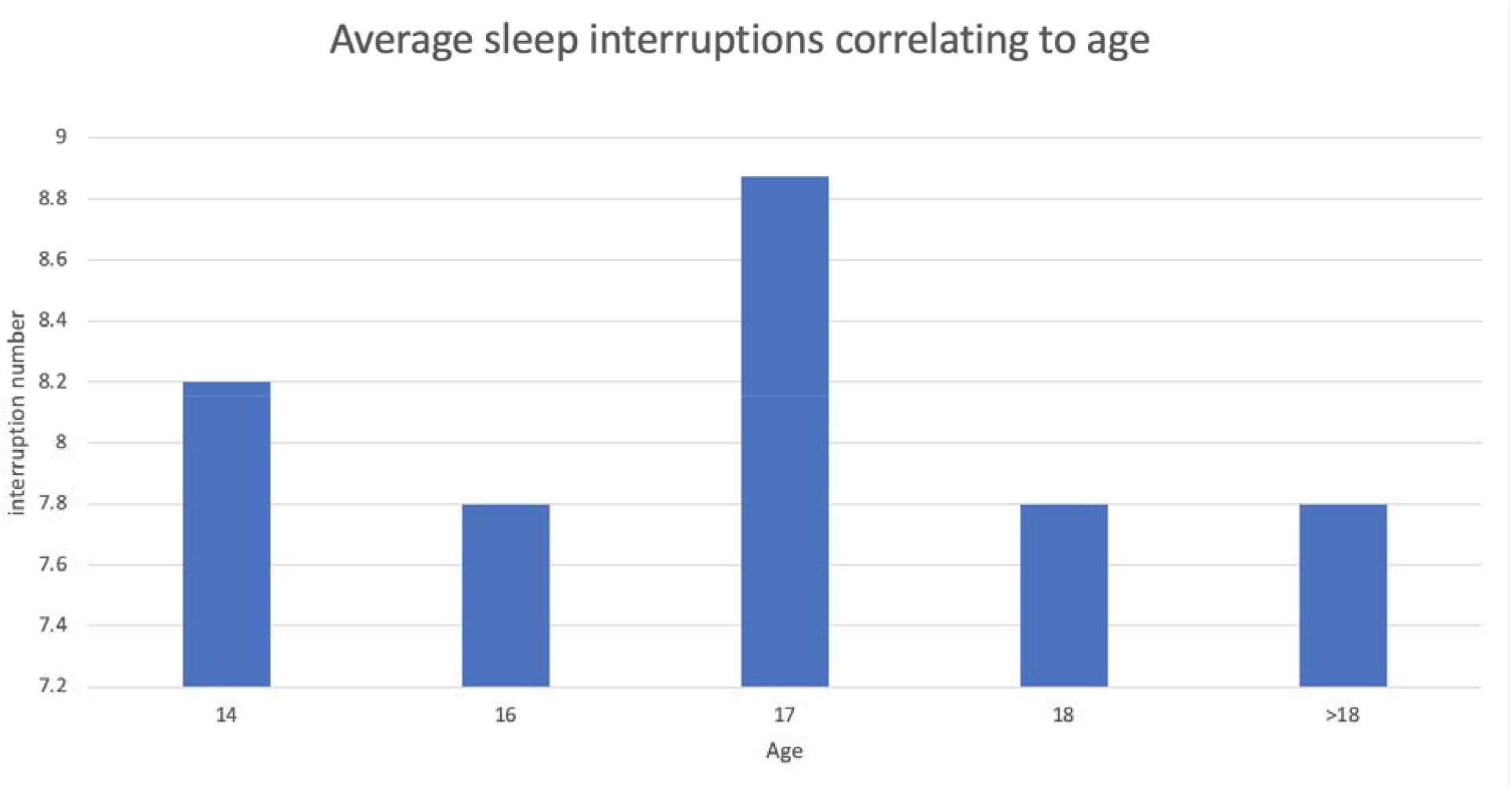
The data in Figure 3 displays interruption number, a figure derived from looking at how many times one woke up during the past month and interruptions to sleep. A greater value of the interruption number correlates with a worse sleep quality. Given that the population was not evenly distributed, the interruption number value was obtained using an arithmetic mean. A general trend may be seen in that sleep interruption number tends to decrease with an increase in age. Given the figures analyzing sleep quality, it may be useful to look into caffeine consumption and the general trends associated with consumption.

**Figure 4.**
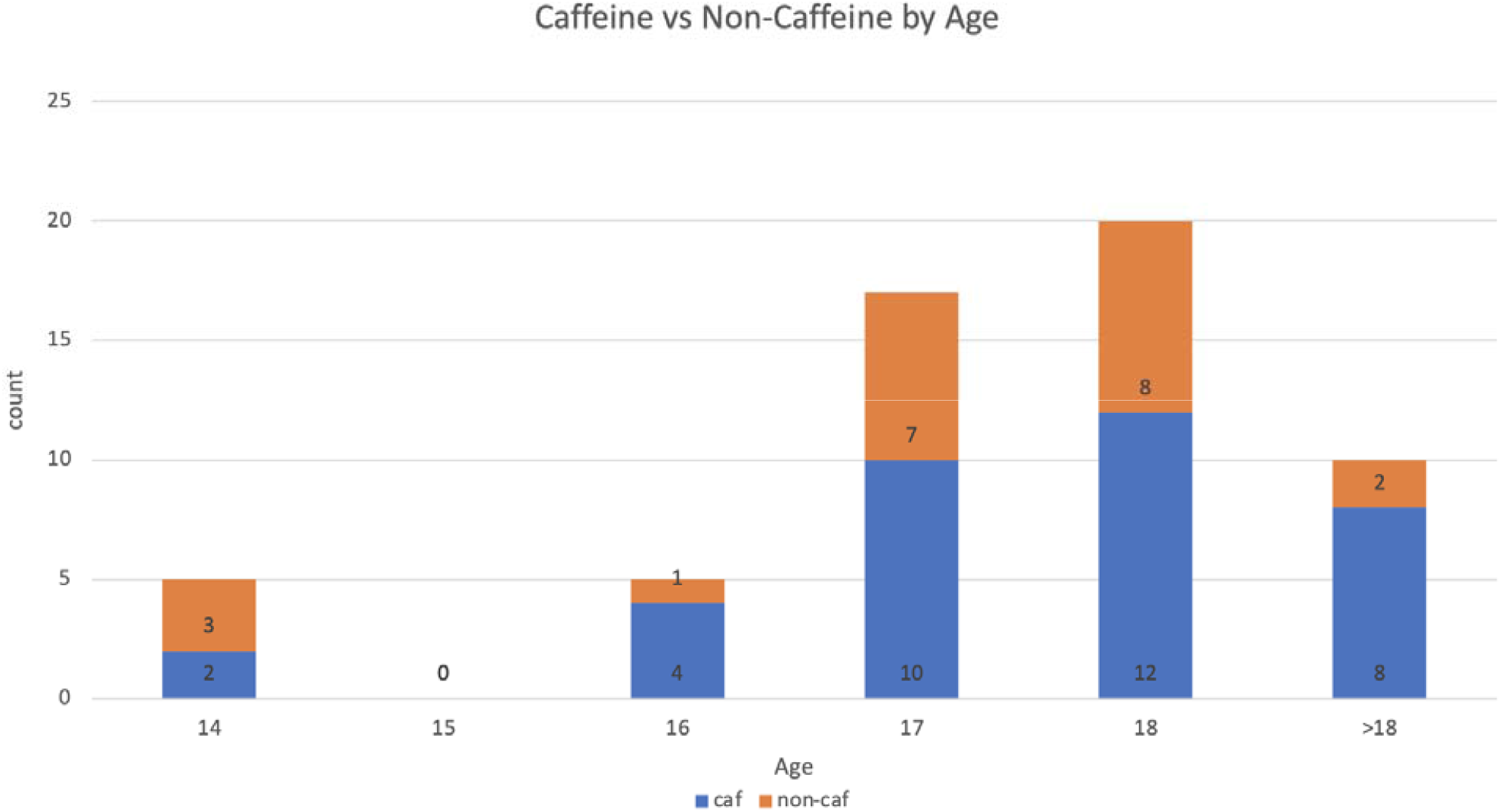
One such display can be seen in sorting caffeine consumption by age. The general trend seems to indicate that the younger groups tend to have less caffeine consumers (14 and 16) as opposed to older age groups. This being said, the caffeine consumption may occur in moderation. As such, it may be useful to look into caffeine intake as an expression in milligrams of caffeine.

**Figure 5.**
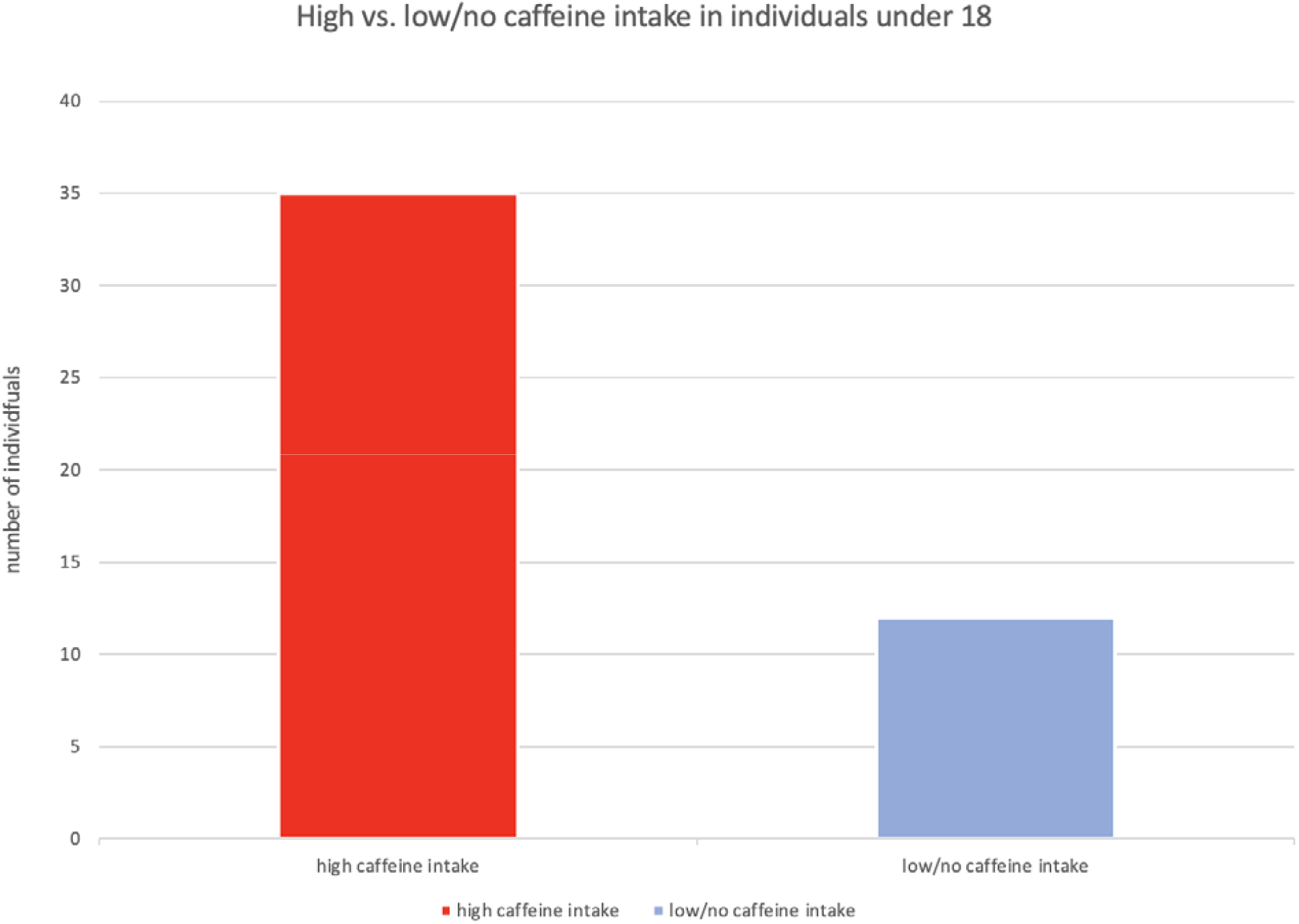
Utilizing the CDC caffeine consumption guidelines for individuals 18 and under, Figure 5 displays that the vast majority of responses exceeded the intake guidelines.

**Figure 6.**
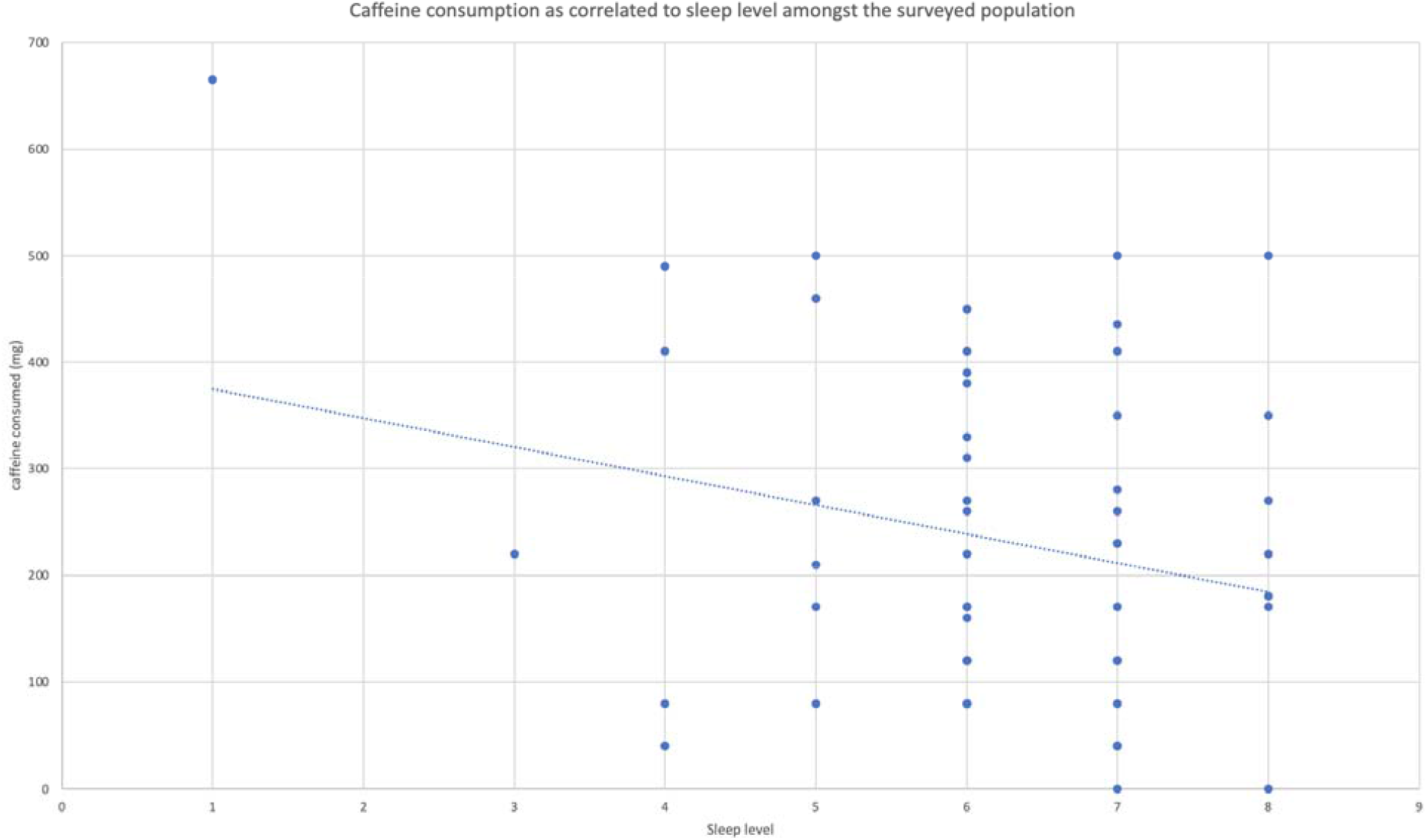
Figure 6 displays caffeine consumption mapped across sleep level for all surveyors. The trend line displays that a low level of sleep correlated with increased caffeine intake. The caffeine consumption in mg expressed was calculated through estimating the caffeine consumption of the caffeine containing beverages/foods reported in the survey. This was done to standardized caffeine consumption. Additionally, existing literature tends to measure, analyze, and report caffeine in mg.

**Figure 7.**
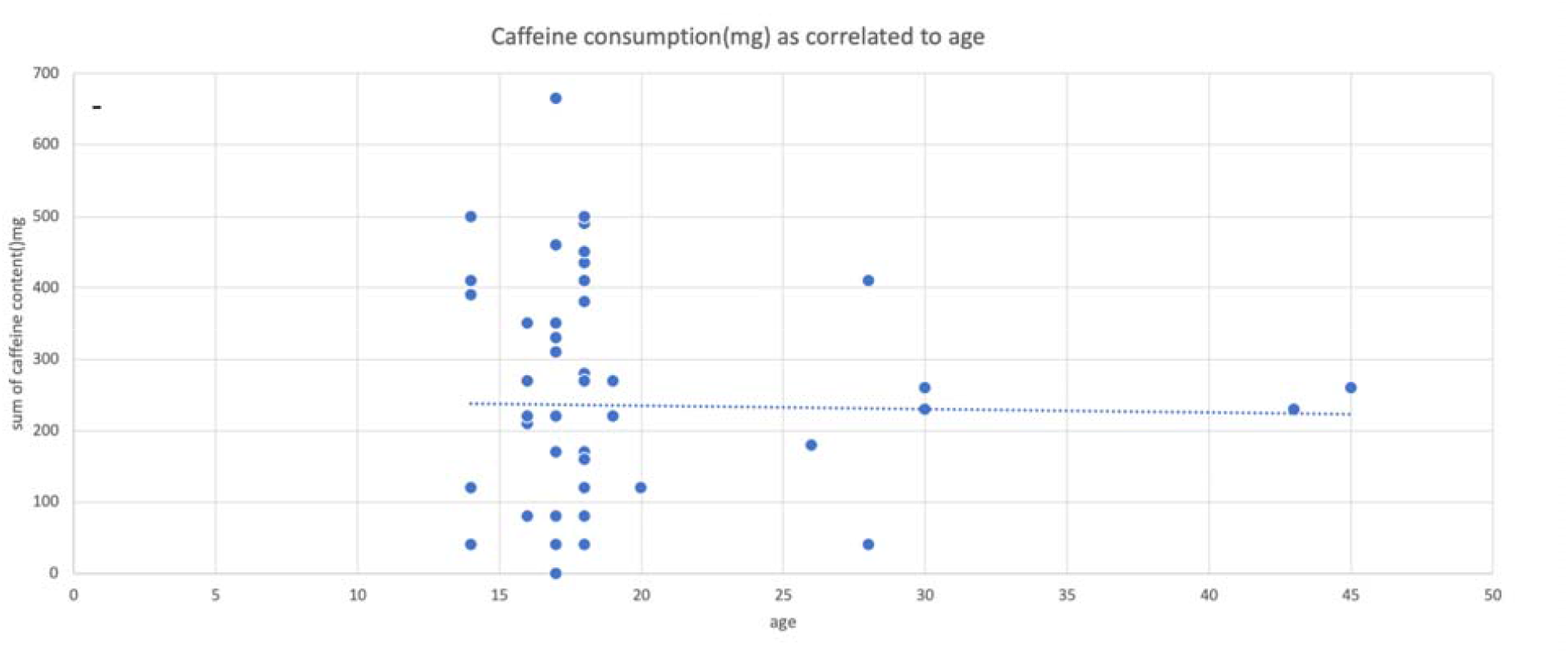
Figure 7 displays the trend between caffeine consumption and age. The general trend expressed by the trendline displays little correlation between the two variables.

**Figure 8.**
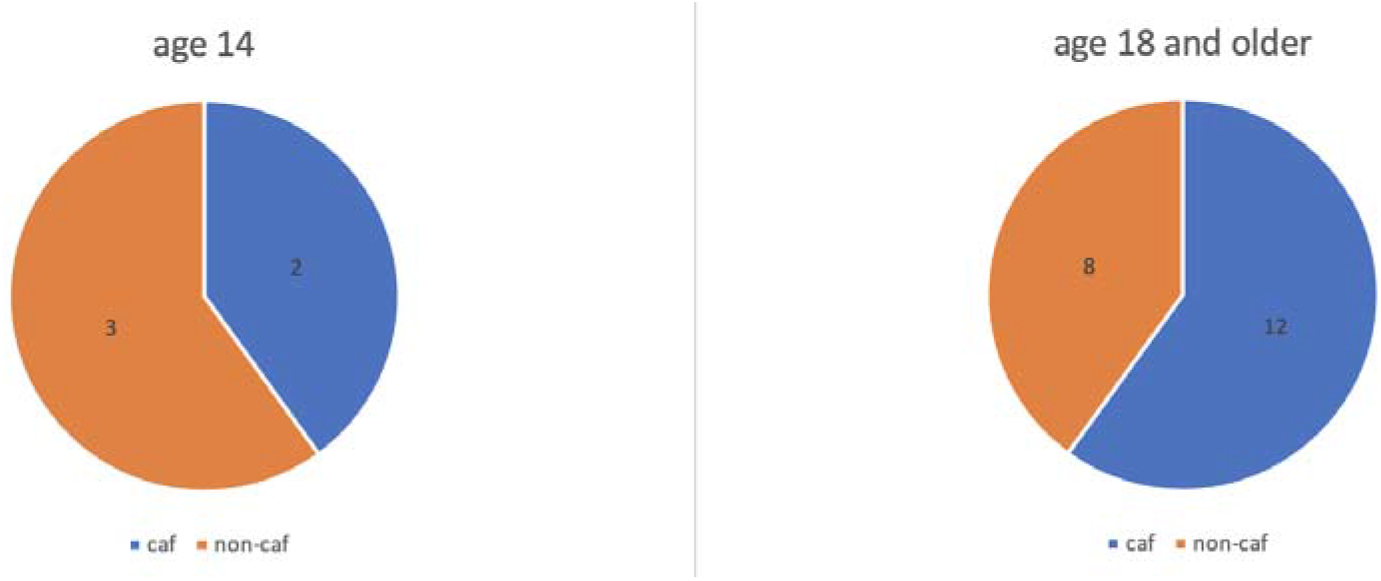
Figure 8 compares the caffeine consumption in the oldest and youngest age groups in the study. There can be comparisons drawn between the difference in the percentage of caffeine consumers in the two age groups.

One important consideration for caffeine consumption is accessibility and onset of caffeine consumption. The 2018 Ágoston et al. publication commented on the motives driving caffeine consumption and the reasoning differences depending on age. Specifically, the article commented on the younger ages wanting to increase as a primary motive for caffeine consumption (Ágoston et al., 2017). In relation to this study, the accessibility factor was likely easily accessible and the onset can be attributed to factors such as desire to increase alertness, as mentioned in the Ágoston, C et al study.

During the survey, the primary finding came from analyzing the association between sleep level and caffeine consumed. The general trend observed displayed a low level of sleep correlating with increased caffeine consumption. This data correlates with the information put forth by the O’Callaghan et al. 2018 publication and the idea of the cycle of evolving caffeine consumption and deteriorating sleep (O’Callaghan et al., 2018). It is important to consider that this particular finding was observed regardless of age. In essence, the correlation between decreased sleep and increased caffeine intake seemed to be relevant across the different age and education levels surveyed.

For individuals 18 and under, the American Academy of Pediatrics recommends healthy caffeine intake of no more than 100mg/day. Using the data from the survey responses, the actual caffeine intake in the population was studied. This data suggested that nearly 75% of the 18 and under population studied consumed caffeine above this recommended guideline. For adults, the Food and Drug Administration (FDA) recommends 400 mg as a safe upper limit for daily caffeine intake (Insight, 2017). Given this increased cushion, only 10% of respondents above the age of 18 fit the criteria for too much caffeine consumption.

Survey respondents were given two separate free-response sections (one for caffeine consumption and one for sleeping habits) to offer information on individualistic factors that contributed to their sleeping habits and their caffeine consumption. These responses were then analyzed to identify similarities among responses but also general trends in responses. For sleep, 15% of respondents indicated that they had been able to get sufficient sleep because of stress. Nearly 27% of respondents indicated that they were unable to sleep due to stress brought on by school or homework which they had to allot additional time to complete. It was interesting to see the recurrence of school, work, and associated stress in the responses. It is important to remember that sleep deprivation, and low levels of sleep in general, is associated with an inability to concentrate and a general decrease in ability to focus (Goel et al, 2009). High levels of caffeine consumption are also associated with symptoms of anxiety, irritability, as well as headaches. These symptoms may be seen as being associated with sleeplessness, stress, as well as feelings of a “racing mind”. All of which may be connected to the sleeplessness and stress indicated by respondents.

## Implications

Potential implications of the study can be seen in improving current schooling to allow for better sleep schedules. Current early school Start times have been seen as leading to sleep deprivation. A 2015 US national comorbidity study published in the American Journal of Public Health discussed the role of early school start times in inadequate sleep. The study investigated the differences in sleep quality between schools with start times before 8 am and those after. The findings displayed clearly poorer sleep quality in students with early start times (Paksarian, 2015). Seeing as caffeine intake was linked with inadequate sleep, and sleep deprivation coupled with caffeine intake was associated with development of caffeine dependence, there is likely a link between school start times and caffeine intake. There may also be directives taken to reduce stress and anxiety in students. One of the biggest findings related to the prevalence of inadequate sleep in the surveyed population. Given the implications of sleep loss, specifically loss of cognitive functionality and increased anxiety and depression symptoms, it is important to prioritize the sleep and associated mental health of students. Additionally, the decreased cognitive functionality speaks to the correlation between sleep deprivation and increased risk of motor vehicular accidents (Hershner & Chervin, 2014). Given that motor vehicular accidents are currently one of the greatest risk factors for adolescents and young adults, it is important to focus on remediation of sleep deprivation, especially among this group (“Risks and Protective Factors”, n.d.).

## Limitations

The students come from the Tri Cities area in the North East Tennessee Area. The largest city in the area Johnson City has a population of just over 66,000 (“U.S. Census Bureau QuickFacts: Johnson City city, Tennessee”, 2021). Thus the sample may not serve as an adequate sampling of the United States or of the entire State of Tennessee. In addition to this, the sample studied was small, making extrapolation of results difficult as well. There are also other confounding factors that may have contributed to loss of sleep as well as increased caffeine usage. Certain examples of this can be seen in use of electronics at night as discussed in the 2009 Calamaro et al publication. There may also have been certain factors that would mean that the actual individualistic healthy caffeine consumption guidelines would be different than those generally set by the FDA. One example of this can be seen in caffeine metabolism. These discrepancies in caffeine metabolism may mean that a healthy maximum level of caffeine consumption may actually be lower than that suggested by the FDA. A 2018 publication from Nehlig discussed that certain genetic and physiological components may affect the rate of caffeine metabolism (Nehlig, 2018). Other similar factors which may alter caffeine metabolism are consumption of other substances, liver disease, diet, oral contraceptives, and many other factors (Langer, 2018). This information seems to indicate that the current 400mg/day safe limit may actually be lower for many individuals, even among those included in this study.

Another major limitation of the study comes from the self reporting survey design. This study design means that respondents indicate what they believe to be true. This would mean that one may believe that they slept more or less than they actually did. Additionally, true quality of sleep may actually differ from the true quality of sleep that occurs. This factor was addressed in the study through including the same question more than once but slightly modified. For example, the respondents were asked how long they slept as well as what time they fell asleep and what time they woke up. The free response section at the end of the survey was meant to help identify potential confounding factors.

## Conclusions

### Sleep

Sleep was the first section studied in the survey distributed. Generally speaking, the survey data was consistent with the statistics offered by the CDC relating to inadequate sleep in citizens of the United States (“How Much Sleep Do I Need?, 2017). In the survey, respondents were asked to provide the approximate number of hours they typically slept in one night. Analysis of the responses indicated that 86% of respondents 18 and under reported sleep levels - with the average sleep between 6 and 7 hours - which were less than the recommended guidelines published by the CDC. The quality of sleep was also analyzed and revealed data concurrent with the inadequate sleep. Though quality of sleep is a difficult measurement through a self-assessment, the results from this section revealed that 25% of those from the 18 and under age group reported bad/very bad sleep quality. For those above the age of 18, 44% reported sleep below the guidelines put forth by the CDC. Here 20% of this group reported sleep quality in the bad/very bad range.

### Caffeine

Current caffeine consumption guidelines are not known to be well established, especially for those under 18. Currently, the FDA has not set an upper level guideline for consumption in children. This being said, current literature and various existing studies have indicated that there is caffeine consumption occurring in the younger age groups. This study confirmed this through the indication that 69% of all individuals studied indicated caffeine consumption. Among those that were 18 and under, 58% of all individuals indicated caffeine consumption. Of this group, among those that had indicated caffeine consumption, 85% consumed caffeine above the recommendations set forward by the American Academy of Pediatrics. For those above 18, this statistic is not as significant. This can be somewhat attributed to the rather high upper boundary of 400 mg/daily of caffeine consumption set forth by the FDA. The results from the survey seem to indicate that there is prevalence of caffeine consumption above recommended guidelines in the population studied.

Future explorations into the field may elect to use a larger sample or even look specifically only at highschoolers or college students. Other work may even look into the specific contributors to caffeine consumption as well as when the consumption of caffeine begins. In this study, there was caffeine consumption shown in even the youngest age group studied. As such, it may be useful for future studies to look at even younger students and when their caffeine consumption begins. Additional studies may even focus on the academic performance of those who consumed caffeine compared to those who did not consume caffeine.

## Data Availability

All data produced in the present study are available upon reasonable request to the authors

## Appendix

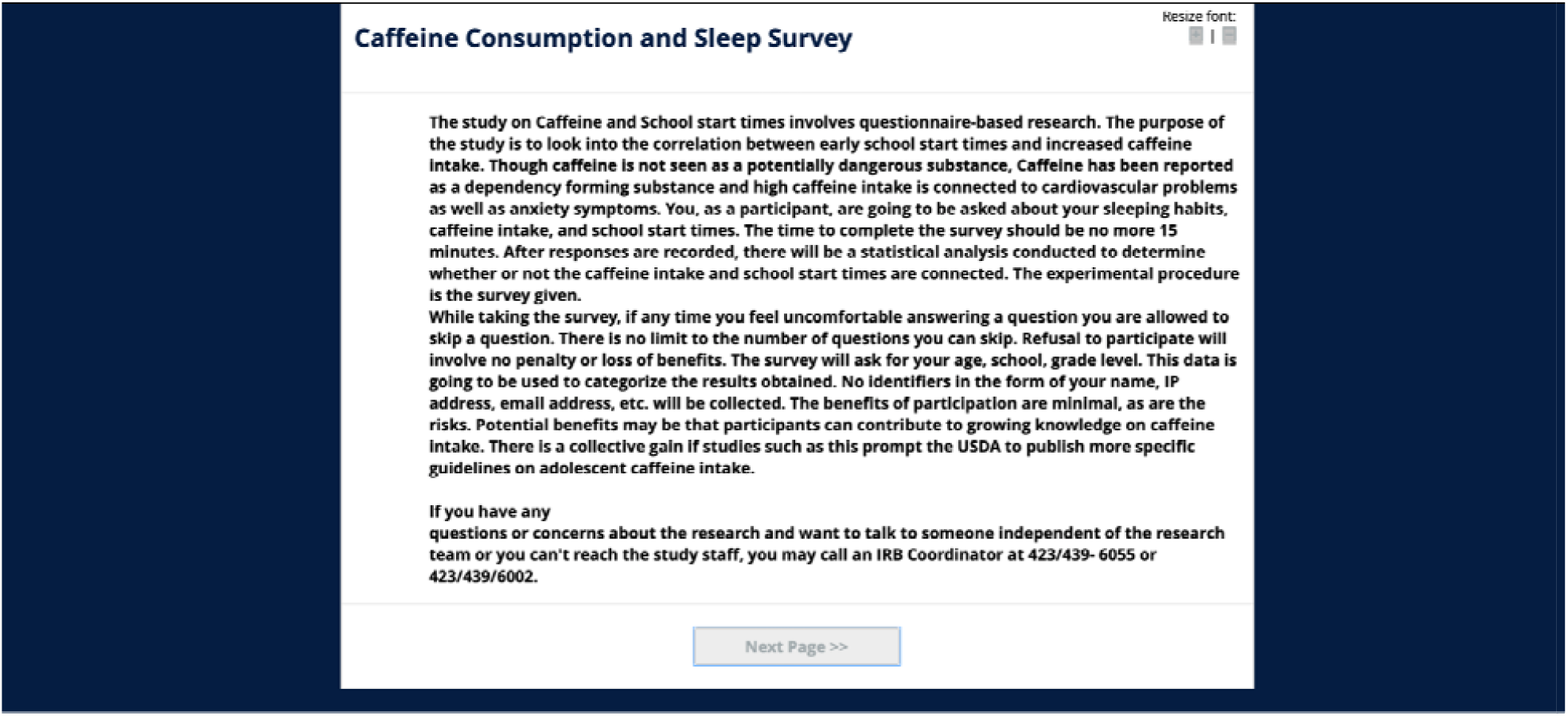

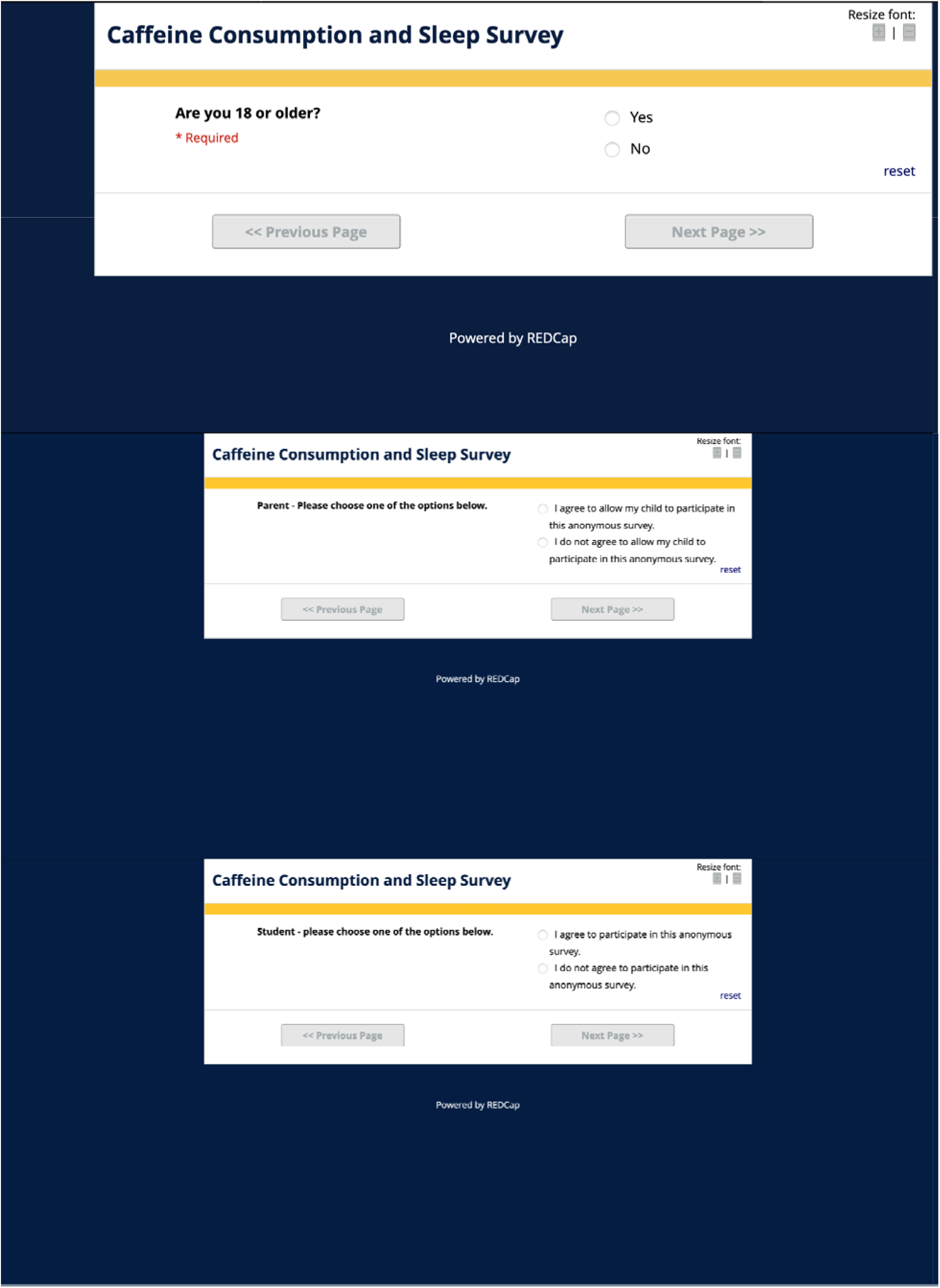

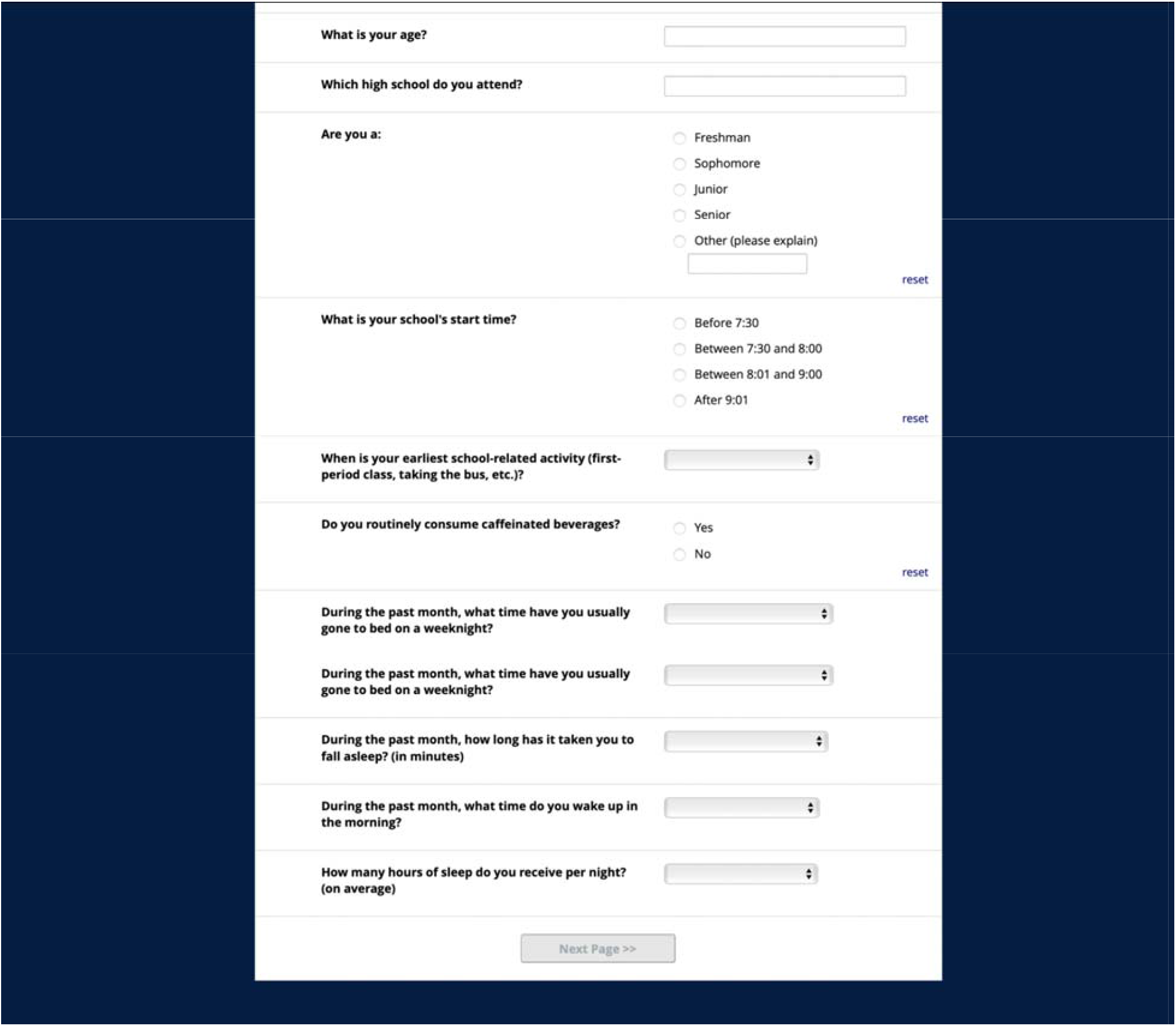

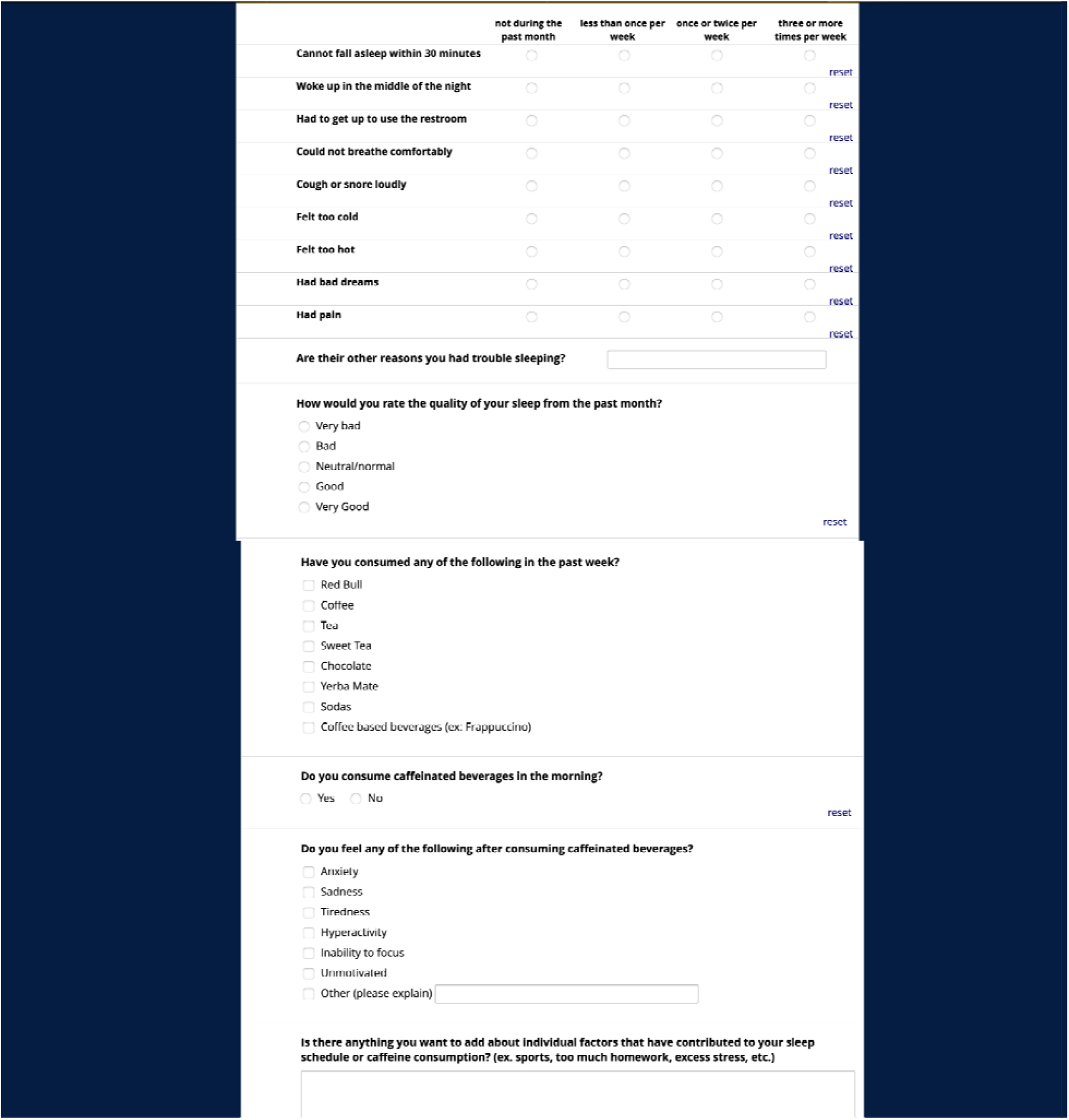

